# Association of socioeconomic and environmental factors with the spatio-temporal variability of suicide rates along a latitudinal gradient

**DOI:** 10.1101/2021.07.06.21260110

**Authors:** Sergio A. Estay, Manuel Ruiz-Aravena, Tomas Baader, Marcelo Gotelli, Cristobal Heskia, Juan Carlos Olivares, Gerardo Rivera

## Abstract

**Background:** Suicide results from complex interactions between biological, psychological, and socioeconomic factors. At the population level, the study of suicide rates and their environmental and social determinants allows us to disentangle some of these complexities and provides support for policy design and preventive actions.

**Objectives:** To evaluate the associations between environmental and socioeconomic factors and demographically stratified suicide rates on large temporal and spatial scales.

**Method:** The dataset contains information about yearly suicides rates by sex and age from 2000 through 2017 along a 4,000 km latitudinal gradient. We used zero-inflated negative binomial models to evaluate the spatio-temporal influence of each environmental and socioeconomic variable on suicide rates at each sex/age combination.

**Results:** Overall, we found differential patterns of associations between suicide rates and explanatory variables by age and sex. Suicide rates in men increases in middle and high latitude regions and intermediate age classes. For adolescent and adult women, we found a similar pattern with an increase in suicide rates at middle and high latitudes. Sex differences measured by the male/female suicide rate ratio shows a marked increase with age. We found that cloudiness has a positive effect on suicide rates in both men and women 24 years old or younger. Regional poverty shows a major impact on men in age classes above 35 years old, an effect that was absent in women. Alcohol and marijuana consumption showed no significant effect sizes.

**Conclusions:** Our findings support high spatio-temporal variability in suicide rates in interaction with extrinsic factors. Several strong differential impacts of environmental and socioeconomic variables on suicide rates depending on sex and age were detected. These results suggest that the design of public policies and interventions to mitigate the impact of the studied variables need to consider the local social and environmental contexts of target populations.

## Introduction

Suicide is one of the leading causes of death globally (Roth et al. 2018), and represents a major concern for health systems. Interventions to prevent or mitigate the risk of suicide are intrinsically challenging since suicide emerges as a result of the complex interactions of biological, psychological and socioeconomic factors at different scales (Jagodic, Agius, and Pregelj 2012). Nowadays, suicide rates show different trends depending on country, age and/or sex (Sinyor, Tse, and Pirkis 2017; Naghavi 2019). Even in this situation, some common demographic patterns have emerged from global data. Men, for instance, present higher suicide rates than women in most countries, and adolescents (10 to 19 years old) show a higher rate of suicide than the rest of the population (Roh, Jung, and Hong 2018). However, beyond these demographic patterns, several others with different trends associated with environmental and socioeconomic factors have been described around the world. At the population level, the spatio-temporal study of suicide rates and their environmental and social determinants allows us to disentangle these complexities and provide support for policy design and preventive actions.

The environmental conditions of places where human populations inhabit have been described as influencing suicide rates in many countries or territories. Among the most cited factors we find sunlight duration, temperature, altitude, and latitude (either by themselves or as proxies of sunlight or temperature), among others. Sunlight or sunshine duration is often proposed as a predictive environmental variable of suicide or suicide rates. Despite the exact biological mechanisms behind this association, several studies have detected statistical associations of sunlight duration, at daily or yearly time scales, and individual suicide events (Vyssoki et al. 2014; Björkstén, Kripke, and Bjerregaard 2009) or suicide rates (Terao et al. 2002; Gao et al. 2019). In the case of altitude or latitude, both variables represent geographic gradients, but they are not real ecological variables, so the interpretation of their significance is always linked to another variable. Most cases where latitude has shown significant association with suicide rates, the interpretation involved its correlation with light-dark cycles or sunlight duration (Lester and Shephard 1998; Voracek and Formann 2004; Lester 1986; Heerlein, Valeria, and Medina 2006; Davis and Lowell 2002; Gao et al. 2019).

Socioeconomic factors, on the other hand, have been more widely analyzed because it has been suggested that they might outweigh climatic factors in explaining regional differences in suicide rates (Jagodic, Agius, and Pregelj 2012; but see Fountoulakis and Gonda 2017). The most studied socioeconomic factors are likely economic deprivation and drug consumption (including alcohol) (e.g., Iemmi et al. 2016; Hasin 2020). Economic deprivation has been considered by using several proxies like GDP, income, changes in employment rates or the occurrence of an economic crisis. Evidence is variable, with marked differences depending on sex and age. The same situation occurs with the spatial scale of the study, which seems to play a key role in the result. At local, country or similar spatial scales most studies pointed to poverty being positively associated with suicide rates (Iemmi et al. 2016; Hsu, Chang, and Yip 2019; but see Bando et al. 2012), although with variable effect size depended on sex. However, at regional or global scale some studies have shown a marginally positive trend between income (or some proxy) and suicide rates (Moniruzzaman and Andersson 2008; Noh 2009). In the case of drug consumption, several studies have shown a link between suicide attempts and the use of drugs (marijuana, inhalants, etc.), especially among adolescents or young people (Hahm et al. 2013;

Carvalho et al. 2019). However, most of this evidence is from studies at the individual level (cases), while information at the population level (rates) is scarcer. Nevertheless, under some circumstances, this link could also be observed at the population level. The prevalence of alcohol use disorder (AUD) has been linked to suicide attempts (Borges and Loera 2010), suicide events (Edwards et al. 2020; Sher 2006), and suicide rates (Razvodovsky 2011). Most studies have pointed to increasing alcohol consumption as a predictor of suicide or suicide rates despite the strength of the association being highly variable. Thus, the relationship between socioeconomic factors and suicide rates seems to be complex and idiosyncratic, with evidence supporting positive and negative associations.

Despite this complex scenario, with variables having different impacts on magnitude and direction, clarifying the role of each factor in the suicide rates and their interaction with demography (e.g., sex and age) is a key component in the design and implementation of preventive policies. The task of simultaneously testing the influence of each of these variables demands not only a multidimensional dataset, but also demands widespread spatio-temporal data coverage. In this study, we evaluate the association between demographically stratified suicide rates on large temporal and spatial scales and environmental and socioeconomic factors. By identifying relevant patterns in suicide rates, this study endeavors to provide insight for policy design and implementation at the local and regional scales.

## Methods

### Data description

#### Suicide rates

Data on suicide events were obtained from the open repository of the Department of Health Statistics and Information (DEIS in Spanish, https://deis.minsal.cl#estadisticas), Ministry of Health, Chile. This dataset contains information about a) the number of suicides by sex and age (5-year categories from 0 to 80+) for each year from 2000 to 2017 in each administrative region of Chile (16 administrative regions), b) total population at each sex/age/region/year combination, and c) age-specific suicide rate for each combination (a/b). We used data from age classes 10-14 to 80+. In total, our dataset involves 32,769 suicide cases, 27,059 in men and 5,710 in women. Due to the geographical location of Chile, the data represents a large latitudinal gradient from 18°S to 53°S (considering the latitude of each regional capital city), equivalent to more than 4,000 km. This latitudinal gradient involves large environmental differences. At 18°S we find the hot desert climate, and at 54°S we find the subpolar oceanic climate according to the Köppen classification.

#### Environmental data

We used two environmental variables related to the availability of sunlight. The first one is total sunlight hours (hereafter Sunlight) in autumn-winter months. This variable is representative of exposure to natural light during the darkest period of the year. Total sunlight hours were calculated using the formula proposed by Forsythe et al. (2003), which uses latitude and day of the year to calculate the number of hours between sunrise and sunset. For calculations we used the latitude of the capital city of each region, where most of the population lives. This definition of total sunlight hours does not consider meteorological conditions, so it represents the maximum amount of sunlight a person potentially experiences at a given place. For this reason, we included a second variable, the proportion of days in a year with significant cloud cover (hereafter Cloudiness) in the capital city of each region. This variable works as a complement to sunlight hours, and as a measure of the net sunlight availability. Cloudiness was obtained from Wilson & Jetz (2016). Together, both Sunlight and Cloudiness offer us an adequate representation of the availability of sunlight along the latitudinal gradient.

#### Socioeconomic data

We included three socioeconomic variables in the analysis. First, we used the percentage of people living with an income below the poverty threshold (hereafter Poverty) as a measure of economic deprivation (as of January 2021 this value is ∼245 USD per person living in Chile per month). Data were obtained for every year from the public repository SINIM (National System of Municipal Information in Spanish, http://datos.sinim.gov.cl). This poverty index is obtained from the CASEN (Socio-Economic Characterization in Spanish) national survey. Each survey can be representative of one or more years depending on the dates. For example, CASEN for 2000 is representative of 2000 and 2001. In addition, we included alcohol and marijuana (cannabis) consumption. In both cases we used the prevalence (%) of consumption in the previous year in the population of each region. Data were obtained from the 13^th^ National Study on Drug Consumption (freely available at http://www.senda.gob.cl/observatorio/). Given that the administrative territory of the Region del Ñuble (∼36° S) was separated from the original Region del Biobio in 2017, no data about drug consumption are available for the current territory of Ñuble. In the past, this territory was included in the reports from the Region del Biobio. Therefore, in this study, historical data were imputed to both regions (Ñuble and Biobio).

### Statistical analyses

As a first step, we estimated general characteristics of our dataset. We evaluated the linear trends of each sex/age/region combination over the period. We used Kendall correlation coefficients for this estimation due to the high non-normality of the data. Additionally, to assess sex differences, we calculated the male/female ratio in suicide rates for each year.

To evaluate the influence of each variable on each sex/age combination, we used a zero-inflated negative binomial model. We chose this kind of model due to the high non-normality of our data because of the high frequency of zeros in some age/sex combinations. In the same vein, our data show a large difference between mean and standard deviation, which makes negative binomial distribution a better choice than the classical Poisson distribution (Gardner, Mulvey, and Shaw 1995; Hilbe 2011; Greene 1994). Suicide rates were modelled by using the number of suicides as a response variable and the logarithm of population size as the offset (McCullagh and Nelder 1989; Madsen and Thyregod 2010; Myers et al. 2012). All variables were standardized by using robust transformation ((x_i_ -Median)/MAD) (Maronna, Martin, and Yohai 2019); thus, all coefficients are comparable because they are at the same scale. The effect size of each variable is shown as the estimated coefficient and its 95% confidence interval (Cumming 2013, 2007). We used McFadden’s pseudo-R^2^ as an approximate metric of goodness of fit for our model formulations. Analyses were performed in the R environment (R Team 2016) and pscl package (Jackman 2020). Codes for analyses are available as supplementary information 1b.

## Results

Overall, suicide rates did not present general temporal trends in the raw data for either sex. Nevertheless, some trends for specific sex, region or age class were detected. For men, an increase in suicide rates was observed in individuals between 30 and 45 years old at mid- (∼30°S) or high- (∼45°S) latitude regions (Fig 1a). For adolescent and adult women (<65 years old), a marked trend in increased suicide rates was observed at mid- (30-35°S) and high- (>40°S) latitudes (Fig 1b). For people in age classes >65 years, only a mild increase trend is noted, mainly in women (Fig 1a-b).

**Figure 1.**
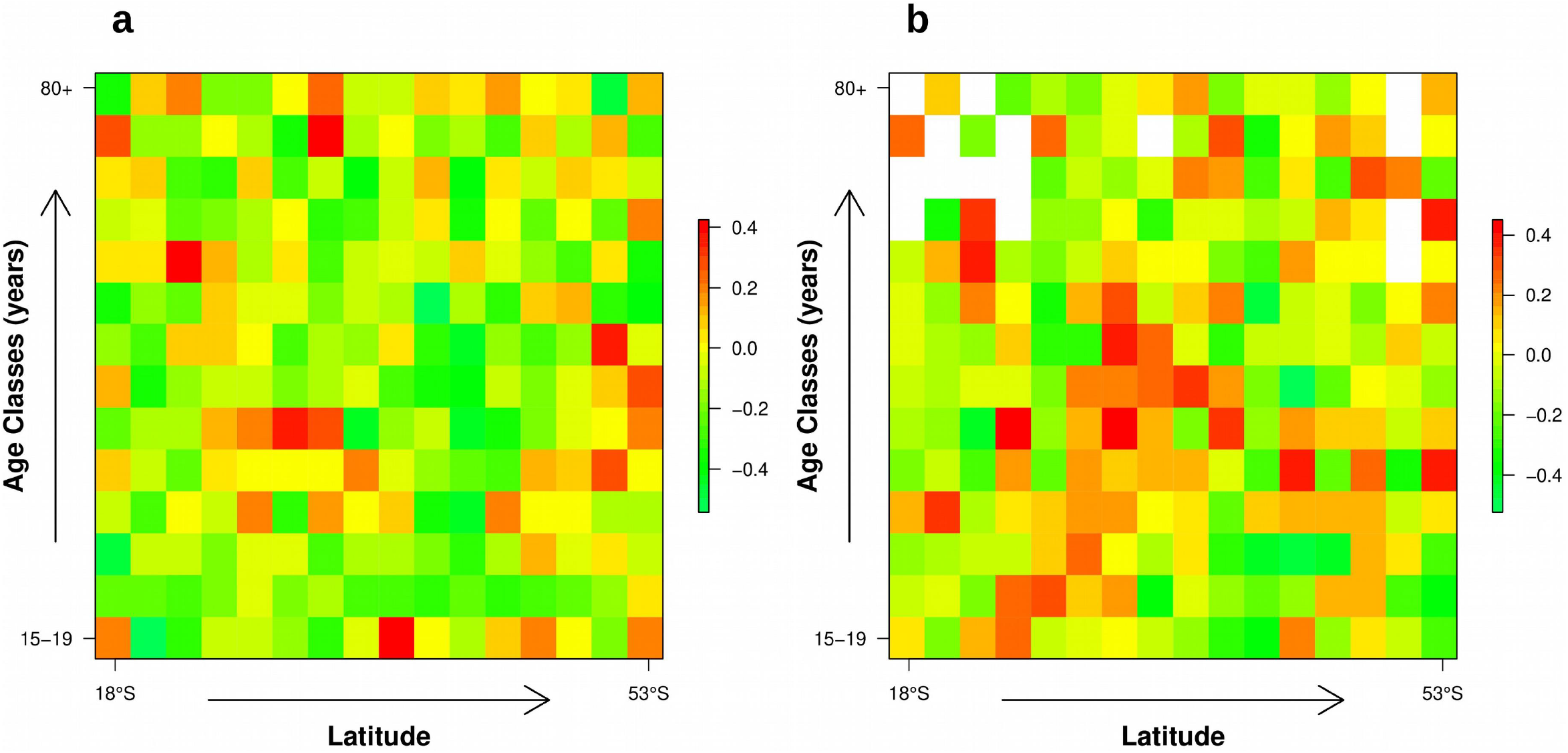
Linear trends in suicide rates through time for men (a) and women (b) by age and latitude estimated using the Kendall correlation coefficient. Empty cells are combinations with too low variability for calculating the coefficient.

From 2000 to 2017 the ratio of male/female suicides ranged from a minimum of 3.85 in 2015 to 6.12 in 2000 (Fig 2a). From 2000 to 2009 the ratio noticeably decreased, but from 2010 it increased. The male/female ratio in adolescents is as low as 1.51 (10-14 years old) and peaks at 8.44 in the 70-74 year age class (Fig 2b). The ratio shows a marked increase with age (Fig 2b) from 10-14 to 20-24-years old. From age class 25-29 to 50-54 the ratio remains almost constant between 4.5 and 5.0. From age class 55-59 the ratio increases again.

**Figure 2.**
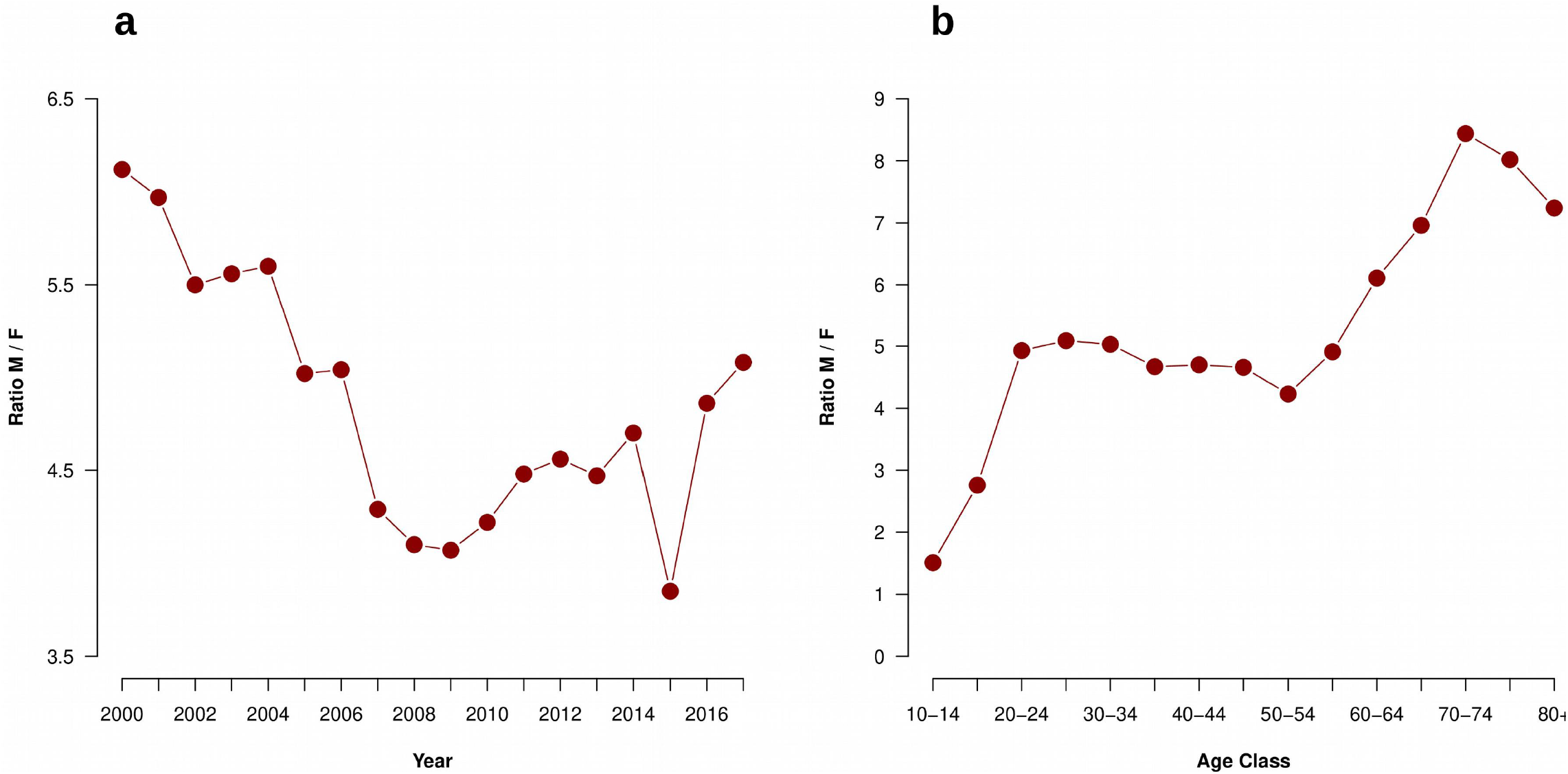
Trends in the male/female ratios of suicide over time (a) and age classes (b).

Analyses on the effect of each variable on suicide rates reveals some interesting patterns. First, the level of Cloudiness has a clear positive effect on suicide rates on men and women 24 years old or younger (Fig 3-4). The pattern was particularly clear for the first three age classes in men and the first two age classes in women (confidence intervals do not contain a zero). For people over 24 years old, the estimated coefficients are mostly positive, as expected, but the previous pattern disappears. In the case of Sunlight, despite the consistently negative effect observed (as expected) in both sexes and age classes, confident intervals contain a zero in almost all cases (Fig 3-4, Suppl. 1a). Regional poverty shows the clearest pattern in the analysis. In the case of men in age classes >35 years, the effect size was markedly positive and increased with age (Fig 3, Suppl. 1a). However, this pattern is absent in women (Fig 4). Alcohol consumption showed no significant effect sizes with the exception of a few sex/age combinations (Fig 3-4, Suppl. 1a). Marijuana use showed an overall weak negative effect, but this effect is not consistent between sexes (Fig 3-4, Suppl. 1a).

**Figure 3.**
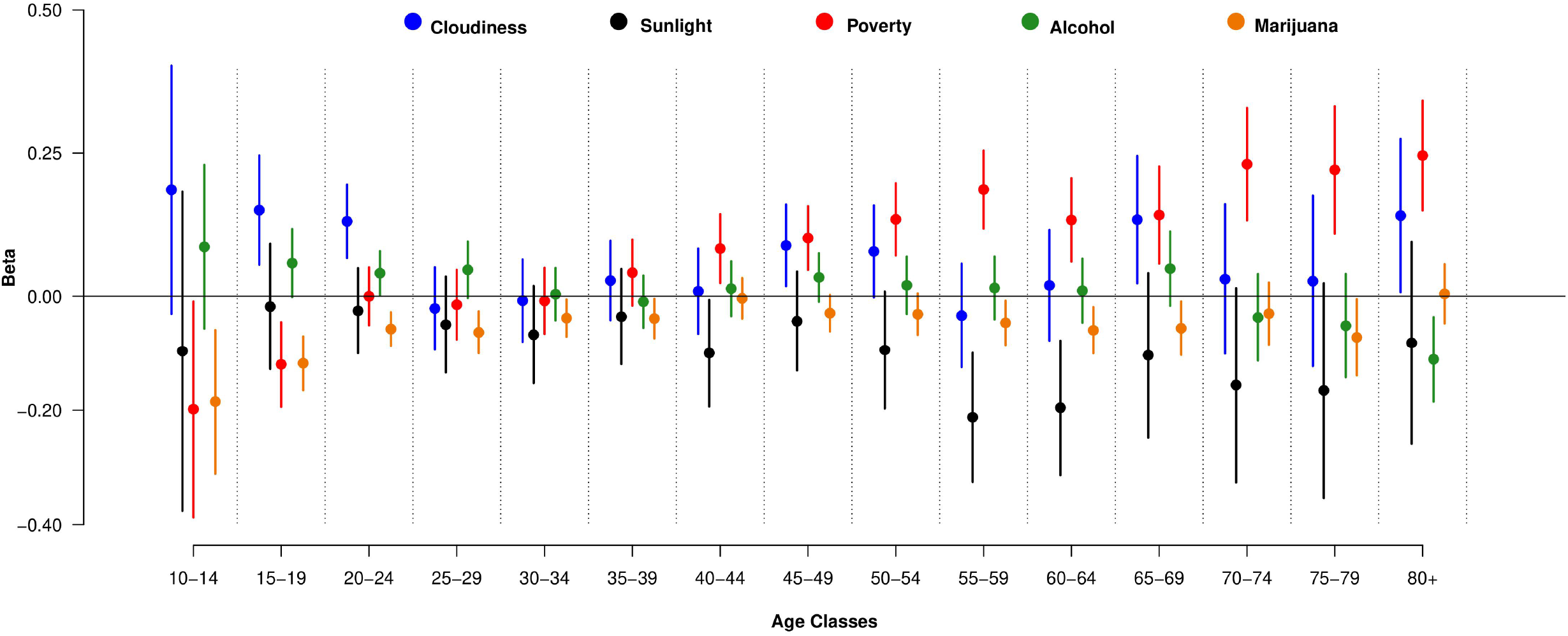
Estimated coefficients and confidence intervals (bars) for the effect of environmental and socioeconomic variables on men by age class.

**Figure 4.**
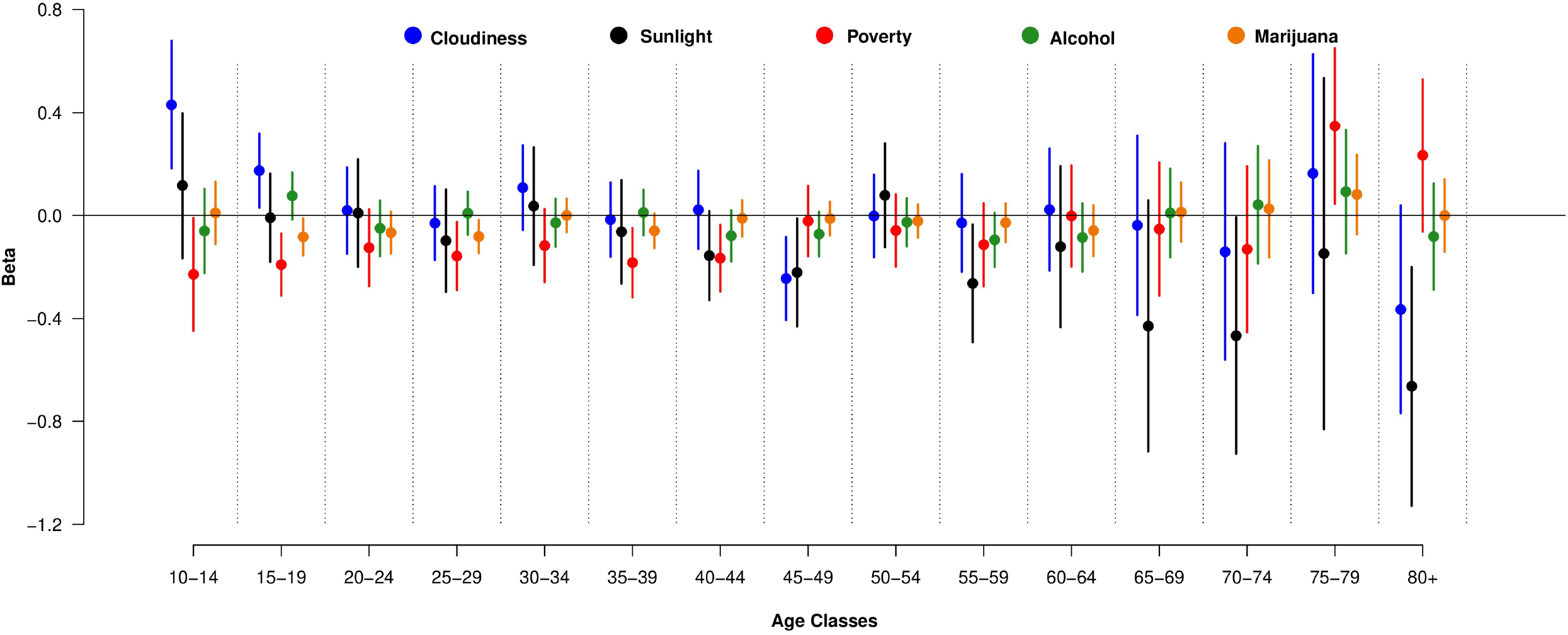
Estimated coefficients and confidence intervals (bars) for the effect of environmental and socioeconomic variables on women by age class.

## Discussion

Our results show a highly idiosyncratic scenario for trends in suicide rates depending on location and age class. In Chile, our results show that the global suicide rate has presented relatively stable levels in the last 18 years. However, some worrying results point to significant local increases in suicide rates in both sexes, especially for people under 45 years of age. The age-standardized suicide rate has decreased significantly in the world (Sinyor, Tse, and Pirkis 2017; Naghavi 2019). The same pattern is observed in Latin America with a clear decrease in recent decades (Naghavi 2019). The major impact of suicide in young adults has been previously described (Naghavi 2019), and it is in this age range where suicide has been consistently ranked among the five leading causes of death (Naghavi 2019; Roth et al. 2018). Similar trends have emerged in other countries, for example in Brazil between 2000 and 2016, an increase in suicide rates was observed in men between 20 and 39 years old, while in women the upward trend was detected in people over 40 years (Martini et al. 2019). In South Korea, on the other hand, a clear increase is noted in men between 30 and 49 years old between 1993-2016, but it is absent below or above these age classes (Lee et al. 2018). In the same country, suicide rates in women 10-19 years old showed a noticeable increase, similar to our case, between 1998 and 2009; however, after this year the increase disappears (Lee et al. 2018). The upward trend in older adults is weak in both sexes and only detectable at middle to high latitudes (Fig 1a-b), which is markedly different to other Latin American countries like Brazil, where this positive trend has been highly significant in the last two decades (Santos et al. 2017).

Sex differences in suicide rates have been well documented with the male/female suicide mortality ratio strongly biased to men (Naghavi 2019); however, the magnitude of this bias is highly variable. In our study, the male/female ratio decreased from 2000 to 2009 and then rose until 2017. Despite the complexity of comparing with other places, a slightly similar pattern (but with lower values for the ratio) was observed in South Korea, with an initial decrease starting around 1993 until 2005-2010, and then an increase in the range 2011-2016 (Lee et al. 2018), something quite different to the almost null change in Brazil (Martini et al. 2019), a Latin American country where more cultural and social commonalities are expected. On the other hand, when we look at the male/female ratio through age classes, the pattern in Chile is substantially different to the global pattern. Globally, the ratio is close to 1 in adolescents and increases to values close to 2.5-3 between people 30 to 60 years old. Over 60 years of age, the global pattern shows a decline in the ratio to values close to 1.5 (Global Burden of Disease Collaborative Network 2019). First, our data show ratio values slightly over 1 in adolescents, which is in line with global patterns, but for middle-aged people we observe higher values for the ratio, with values ranging between 4 and 5 (Fig 2b), which is 30 to 50% higher than global data. The difference is even clearer in people over 60 years old, where ratios rise to values above 8 (Fig 2b). However, ratio values over 4 are not uncommon in Latin America or some countries in Asia (Global Burden of Disease Collaborative Network 2019), and very high ratios in older adults have been previously reported in Brazil (Martini et al. 2019). We will discuss some potential explanations for these trends in the next paragraphs.

The evaluation of environmental and socioeconomic variables showed two main patterns. The first is the positive influence of cloudiness as a metric of the net sunlight available on suicide rates in both sexes under 25 years old. This particular result was, in some sense, backed up by the consistent, although non-significant, negative estimations of the sunlight coefficients. The latitudinal pattern of cloudiness (and sunlight) is remarkable in Chile, with almost cloud-free conditions in northern regions to clouds covering the skies in southern regions for more than 65% of the year (Wilson and Jetz 2016). The association of sunlight availability (or some proxy) with suicide rates or depression has been described many times in previous studies (Olders 2003; Vyssoki et al. 2014; Hernández, Hernández-Sánchez, and Flores-Gutiérrez 2018; Makris et al. 2021), which supports our results. In fact, studies emphasizing the relationship between suicide rates and latitude often offer explanations using sunlight availability at some temporal scale as the actual variable behind the pattern (Lester and Shephard 1998; Voracek and Formann 2004; Heerlein, Valeria, and Medina 2006; Davis and Lowell 2002). Despite the difficulties of inferring mechanisms from statistical associations, a common explanation for this relationship is that Cloudiness/Sunlight interacts with the biological mechanisms behind the regulation of serotonin and melatonin production, which, in turn, regulate several behaviors linked to suicidal behavior (Vyssoki et al. 2014; White et al. 2015). However, these explanations should be treated with caution because, beyond the consistent statistical pattern described here and other studies, by accepting these explanations we are downscaling population results to the individual level. The influence of cloudiness disappears in people over 25 years old. Estimated coefficients for sunlight were consistently negative, as expected, but non-significant. This result suggests a hidden interaction between age and sunlight availability which mediates suicide rates. As a hypothesis, we can speculate that the influence of being exposed to sunlight is higher on the young population, still under psychological development, than on adults, and that this differential influence is reflected in latitudinal patterns of suicide rates.

The second pattern was even clearer. Our results showed a strong positive influence of poverty on suicide rates in men over 35 years old. The poverty coefficients estimated for all age classes over 35 years were highly (and increasingly) significant. The link between poverty and high suicide rates has been widely described in North America (Fontanella et al. 2018), Asia (Lee et al. 2018), and Europe (Fountoulakis et al. 2014, 2016). The impact of poverty on health conditions, particularly mental health, is so ubiquitous because its detrimental influence acts via many paths. In an ecological interpretation, chronic poverty is a press disturbance, a constant damaging force with a cumulative effect on people’s health, which in turn could be reflected in high chronic suicide rates. Another path is the effect of economic crisis on mental health (Álvaro-Meca et al. 2013; Chang et al. 2013). In this case, economic crises act as pulse disturbances, which could be reflected in sudden increases in suicide rates with short- or long-term impacts depending on the pace of economic recovery. Previous studies have pointed out the effect of poverty or deprivation on suicide rates using unemployment as a proxy. Unemployment is a representative measure of population well-being and, as such, is a useful representation of deprivation. In this regard, evidence shows sporadic increases both during or immediately after economic crises (a sign over time, (Álvaro-Meca et al. 2013; Chang et al. 2013), and a cluster of high suicide rates in localities with high chronic unemployment (a sign over space Trgovac, Kedron, and Bagchi-Sen 2015; Alarcão et al. 2020).

The association between poverty and suicide rates was only observed in men. This situation is similar to the results of Fountoulakis et al. (Fountoulakis et al. 2016). These authors, using unemployment as a measure of economic distress, found that this variable was only important in models for men but not women across Europe. The reason(s) behind these differential results are difficult to evaluate and there is quite likely a complex interaction between biological and cultural factors.

A surprising result was the null effect of alcohol and a weak and inconsistent effect of marijuana on suicide rates. Both have been positively associated with suicide rates (O’Neill and O’Connor 2020; Hasin 2020; Razvodovsky 2011; Sher 2006; Edwards et al. 2020). One possible explanation is that the impact of these factors can be better detected at individual scale, but it disappears on a larger scale, although this is only speculation. The effect of marijuana was negative, weak and inconsistent between sexes, which suggests a spurious effect produced by the large increase in marijuana consumption in the last few decades. In Chile, consumption prevalence has increased from 5.7% in 2000 to 12.7% in 2018, peaking at 14.5% in 2016 (SENDA 2019), which is likely the reason behind this result. Another reason for these results is the type of variables included in this study. We used yearly prevalence of consumption, but prevalence of alcohol and drug use disorder in the population could be a better alternative. Unfortunately, we do not have data on these variables for our time window, which prevents their use in this study. Under this scenario, the interpretation of the lack of effect of alcohol and cannabis consumption in this study should be cautionary.

Our study analyzed suicide rates over a large geographical scale with an ecological approach in the analysis. In this sense, we cannot downscale our conclusions to the individual level. In individuals, promoting and protective factors can interact in ways that can be difficult to detect using population-level data. However, ecological studies like this and several others (e.g., Lee et al. 2018; Sinyor, Tse, and Pirkis 2017; Trgovac, Kedron, and Bagchi-Sen 2015; Dixon et al. 2014) are relevant for designing public policies and interventions to mitigate the impact of the studied variables (Sinyor, Tse, and Pirkis 2017; Steelesmith et al. 2019; World Health Organization 2014). For populations living at high latitudes, where sunlight is scarce in winter or localities with high levels of cloudiness, the implementation of preventive actions specially designed for young people, including improved access to medical services, could be an advantageous strategy. Also, our results point out to potential advantages of interventions specially designed for adult men in chronically deprived populations and/or rapid response programs during and after economic crises (see similar conclusions in Steelesmith et al. 2019; Sinyor, Tse, and Pirkis 2017; Iemmi et al. 2016). Our results emphasize the design of particular preventive approaches depending on sex, age class, and geographic locations. General strategies can be useful on some level, but to take into account other contextual variables, like those described in this study, could be a significant improvement for the success of preventive actions. These interventions should include psychological and social support, and the necessary financial resources, for at-risk populations. We hope our results will be useful for decision-makers.

## Data Availability

Available upon request after acceptance in a journal.

## Acknowledgments

SAE thanks to ANID PIA/BASAL FB0002 for financial support.

---

**Supplementary information 1a.**
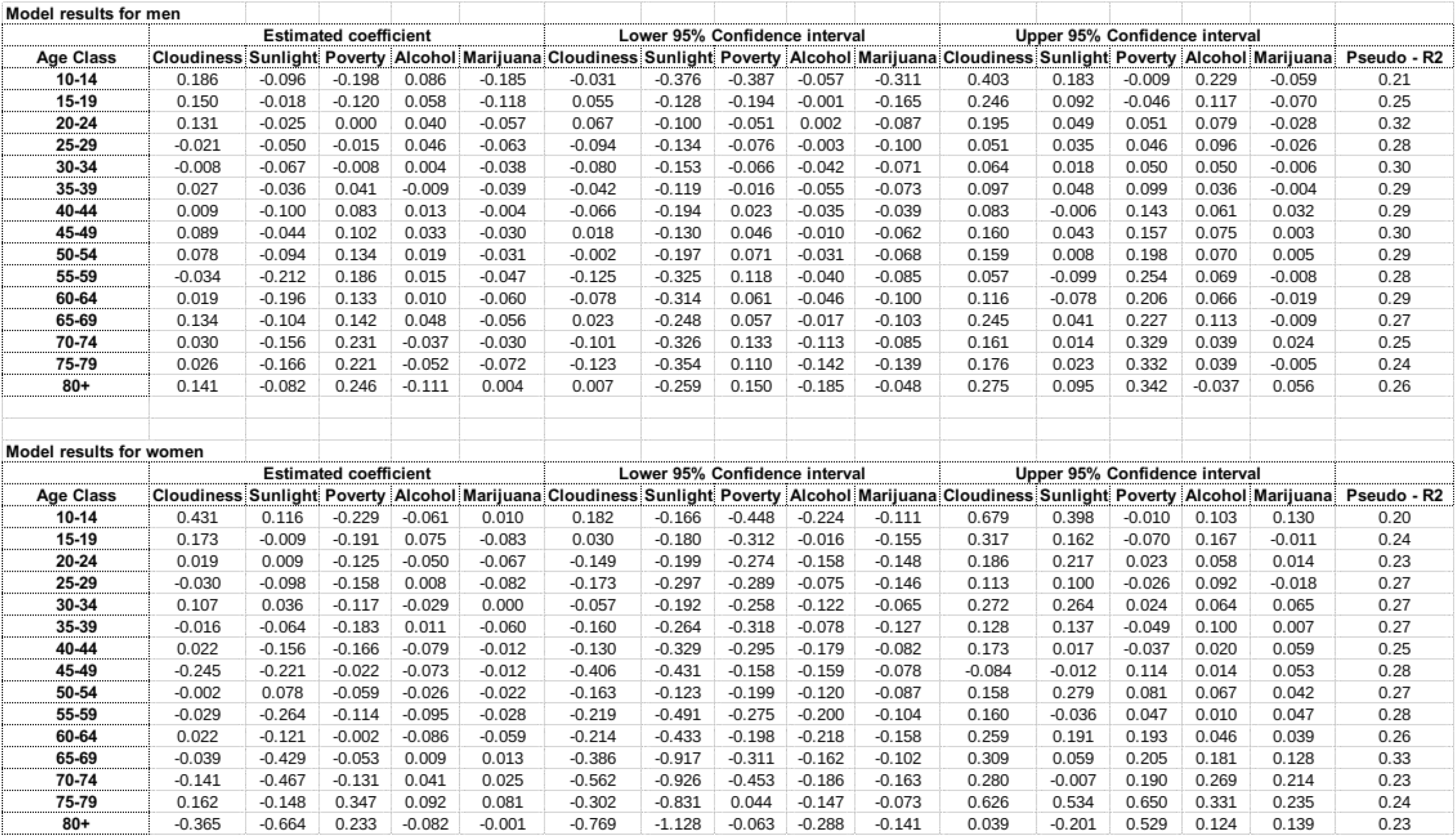

